# Presence of Symptoms 6 Weeks After COVID-19 Among Vaccinated and Unvaccinated U.S. Healthcare Personnel

**DOI:** 10.1101/2022.02.16.22271092

**Authors:** Nicholas M. Mohr, Ian D. Plumb, Karisa K. Harland, Tamara Pilishvili, Katherine E. Fleming-Dutra, Anusha Krishnadasan, Karin F. Hoth, Sharon H. Saydah, Zachary Mankoff, John P. Haran, Melissa Briggs-Hagen, Eliezer Santos León, David A. Talan, the Project PREVENT Network

## Abstract

**Importance:** Although COVID-19 vaccines protect against infection and severe disease, the role of vaccination in preventing prolonged symptoms in those with subsequent infection is unclear.

**Objective:** To determine differences in symptoms stratified by prior vaccination reported by healthcare personnel (HCP) 6 weeks after onset of COVID-19, and whether there were differences in timing of return to work.

**Design:** Nested cohort study within a multicenter vaccine effectiveness study. HCP with COVID-19 between December 2020 and August 2021 were followed up 6 weeks after illness onset.

**Setting:** Health systems in 12 U.S. states.

**Participants:** HCP participating in a vaccine effectiveness study were eligible for inclusion if they had confirmed COVID-19 with either verified mRNA vaccination (symptom onset ≥14 days after two doses) or no prior COVID-19 vaccination. Among 681 eligible participants, 419 (61%) completed a follow-up survey approximately 6 weeks after illness onset.

**Exposures:** Two doses of a COVID-19 mRNA vaccine compared with no COVID-19 vaccine.

**Main outcomes and measures:** Presence of symptoms 6 weeks after onset of COVID-19 illness and days to return to work after COVID-19 illness.

**Results:** Among 419 HCP with confirmed COVID-19, 298 (71%) reported one or more COVID-like symptoms 6 weeks after illness onset, with a lower prevalence among vaccinated participants (60.6%) compared with unvaccinated participants (60.6% vs. 79.1%; aRR 0.70, 95% CI 0.58-0.84). Vaccinated HCP returned to work a median 2.0 days (95% CI 1.0–3.0) sooner than unvaccinated HCP (aHR 1.37; 95% CI, 1.04–1.79).

**Conclusions:** A history of two doses of COVID-19 mRNA vaccine among HCP with COVID-19 illness was associated with decreased risk of COVID-like symptoms at 6 weeks and earlier to return to work. Vaccination is associated with improved recovery from COVID-19, in addition to preventing symptomatic infection.

**KEY POINTS:** *Question:* Does vaccination lead to improved recovery of symptoms and return to work following COVID-19?

*Findings:* In this nested cohort study of healthcare personnel, participants with COVID-19 who had received two doses of a COVID-19 mRNA vaccine were less likely to report symptoms 6 weeks after illness onset than participants with COVID-19 who were unvaccinated. Return to work was sooner if previously vaccinated.

*Meaning:* Vaccination is associated with improved recovery from COVID-19, in addition to prevention of infection and disease.

## Introduction

Since December 2020, two approved messenger RNA (mRNA) vaccines, BNT162b2 (Pfizer-BioNTech) and mRNA-1273 (Moderna), have been available in the United States to prevent severe acute respiratory syndrome coronavirus 2 (SARS-CoV-2) infection, hospitalization, and death.^1-4^ Prolonged and debilitating symptoms associated with COVID-19 have been widely recognized in a proportion of survivors.^5^ Prolonged symptoms occur in both hospitalized and non-hospitalized COVID-19 patients.^6^ Commonly reported symptoms include fatigue, dyspnea, and neurocognitive deficits. Ongoing symptoms can impair survivors’ ability to function and work.^7,8^ The prevalence of these symptoms, however, varies significantly among reports, and the mechanisms underlying these symptoms have not been elucidated.^9-14^

Vaccination may prevent prolonged COVID-19 symptoms through several mechanisms: preventing COVID-19 infection, limiting the severity of acute illness through vaccine-mediated immunity, and affecting the ongoing immune response even after acute infection. In a case series of 39 infections among healthcare workers vaccinated with two doses, symptoms at 6 weeks were common.^15^ A separate study of self-reported vaccination status and symptoms among the general public found that COVID-19 symptoms 4 weeks after infection were less prevalent among the vaccinated.^16^ An analysis of electronic medical record data suggested that vaccination might also prevent other post-acute sequelae such as cardiovascular events, coagulation disorders, pulmonary disorders, and others that comprised a comprehensive risk of symptoms associated with prolonged recovery.^17^

In this study our primary objective was to measure the association between prior mRNA COVID-19 vaccination and symptoms 6 weeks after initial COVID-19 illness among healthcare personnel (HCP). We hypothesized that symptoms would be less common after 6 weeks among the vaccinated because the initial illness among vaccine breakthrough cases is less often severe, and severity of illness is one predictor of the likelihood of prolonged symptoms.^14,18^ We conducted a secondary analysis to evaluate recovery from COVID-19 by assessing whether it took longer to return to work if unvaccinated.

## Methods

### Study Design, Data Collection, and Population

As part of the PReventing Emerging Infections through Vaccine EffectiveNess Testing Project (Project PREVENT), we enrolled HCP who were working on-site at participating academic medical centers who had been tested for SARS-CoV-2. Characteristics of the 15 participating sites are summarized in **Supplemental Table S1**, and details of study protocols and forms for the parent study are available online.^19^ Vaccine coverage reached 84% (interquartile range [IQR] 73% to 91%) at participating sites by September 2021. Data from PREVENT sites and other platforms contributed to vaccine effectiveness analyses that have been reported previously.^3,20^

For this nested cohort study, we included HCP with symptomatic SARS-CoV-2 infection (COVID-19) if they had a positive SARS-CoV-2 nucleic acid amplification test or antigen test and had consistent symptoms (as listed in **Supplemental Table S2**) within 14 days before or after the positive test. Participants provided data by electronic surveys or interviews (online, by phone, or in person). Each participant completed an enrollment survey 14 to 60 days after his/her positive test and was offered a follow-up survey 6 weeks (42 days) after symptom onset (or at the time of enrollment if later than 42 days). We excluded participants from the analysis who had partial vaccination, had received a non-mRNA COVID-19 vaccine, did not have available vaccination records, did not complete the baseline survey within 60 days, or did not complete the follow-up survey by 10 weeks after symptom onset. No participants had received more than 2 vaccine doses.

This activity was reviewed by CDC and was conducted consistent with applicable federal law and CDC policy.^21^ All participants provided informed consent prior to participation. This report satisfies the Strengthening the Reporting of Observational Studies in Epidemiology (STROBE) criteria (Supplemental Table S8).^22^

### Definitions and Data Collection

The 6-week follow-up survey included questions on a variety of symptoms which we categorized into three overlapping groups. We considered *COVID-like symptoms* to be fever, cough, shortness of breath, chills, fatigue, joint pains or muscle aches, headache, loss of taste or smell, sore throat, sinus congestion, diarrhea, nausea, or vomiting.^23,24^ We defined *neurologic symptoms* based on potential etiology as dizziness, headache, muscle weakness, movement problems, confusion, memory difficulties, concentration problems, or loss of taste or smell.^12,25,26^ We defined *any 6-week symptoms* as the symptoms listed above or others included in the 6-week survey: trouble sleeping, exercise problems, chest pain, or abdominal pain. For each symptom included in the 6-week survey, participants were asked to rate perceived severity as mild, moderate, or severe.

The study team verified vaccine status and testing results via occupational health clinics, vaccine cards, state registries, or medical records. We considered participants to be unvaccinated if they had not received any COVID-19 vaccine doses and vaccinated if they had received a second dose of a SARS-CoV-2 mRNA vaccine ≥14 days prior to the positive test. We considered comorbidities to be present at the time of infection if reported on the survey or identified in medical records from the period of acute illness. We classified participants as having two or more comorbidities if they had at least two diagnoses from our predefined list including cardiopulmonary, immunologic, and mental health-related comorbidities (full list available in **Supplemental Table S2**). Several of these conditions have been identified both as risk factors for severe COVID-19 outcomes and as possible risk factors for long term symptoms following COVID-19 illness.^27,28^

As part of the follow-up survey, we asked participants to report the dates when they resumed work. We calculated time to return to work as the number of days from onset of symptoms until the first day at work after illness.

### Statistical Analysis

We defined our primary outcome as the presence of COVID-like symptoms at the time of the 6-week follow-up survey. We conducted additional analyses for neurologic symptoms and for any 6-week symptoms, and we assessed whether symptoms at 6 weeks were also present within 14 days of the date of the positive test. For assessment of all symptom groups, we performed a sensitivity analysis restricted to symptoms rated by participants as “moderate” or “severe.”

We used multivariable Poisson regression with a sandwich variance estimator to model the relative risk of having symptoms at the 6-week follow-up for complete vaccination compared with no vaccination.^29^ In the multivariable model we included categorical variables of age, race and ethnicity, and co-morbidities selected a priori. Comorbidities were represented in a dichotomous variable of two or more comorbidities at baseline to indicate whether chronic illness was present.^30^ We included categorical variables for calendar month of illness and number of weeks from symptom onset and follow-up survey completion to account for temporal changes in the prevalence of symptoms. We also calculated the adjusted risk difference as the difference in proportions of participants reporting symptoms in the follow-up survey by vaccination status, using Poisson regression.^31^

We compared median differences in time to return to work by vaccination status using the Wilcoxon rank sum test. To compare the rate of return to work by vaccination status we constructed Kaplan-Meier survival curves and used the log-rank test. We used a Cox proportional hazards model to calculate an adjusted hazard ratio (aHR) to compare time to return to work between vaccinated and unvaccinated participants, counting zero days if there were no days off work after symptom onset. We included the same covariates as we did in our multivariable Poisson regression, except for time to follow-up survey response. We assessed Schoenfeld residuals to ensure that the proportional hazards assumption was met.

We compared individual symptoms on the follow-up survey between vaccinated and unvaccinated cohorts, using unadjusted prevalence, relative risk, and risk difference (defined as the prevalence in vaccinated minus the prevalence in unvaccinated participants).^32^

## Results

Among 1012 HCP with confirmed COVID-19, 331 were excluded because they were partially vaccinated, they received a non-mRNA vaccine, or vaccination records were unavailable. Among the remaining 681 HCP, 419 (61.5%) completed the baseline survey by 60 days and the follow-up survey between 42 and 70 days after symptom onset. Those who did not complete the follow-up survey had fewer comorbidities and were less likely to be White non-Hispanic (**Supplemental Table S4**). We included 419 HCP with symptom onset between December 28, 2020 and August 26, 2021: 180 (43.0%) who were vaccinated with two doses of an mRNA vaccine and 239 (57.0%) who were not vaccinated (**Figure 1**). Among vaccinated participants there was a median of 24.1 weeks (IQR 15.3–28.1 weeks) between the 2^nd^ vaccine dose and the date of illness onset. Most vaccinated participants received the Pfizer-BioNTech vaccine (n=158, 87.8%); 22 (12.2%) received the Moderna vaccine. Vaccination status varied by race/ethnicity and education (**Table 1, Supplemental Table S5**). Among the 419 participants included in the analysis, 260 (62.1%) participants provided direct clinical care, and 296 (70.6%) worked in acute care hospitals. Only 1 participant (0.2%) required hospital admission for acute COVID-19, and no participants died.

**Figure 1.**
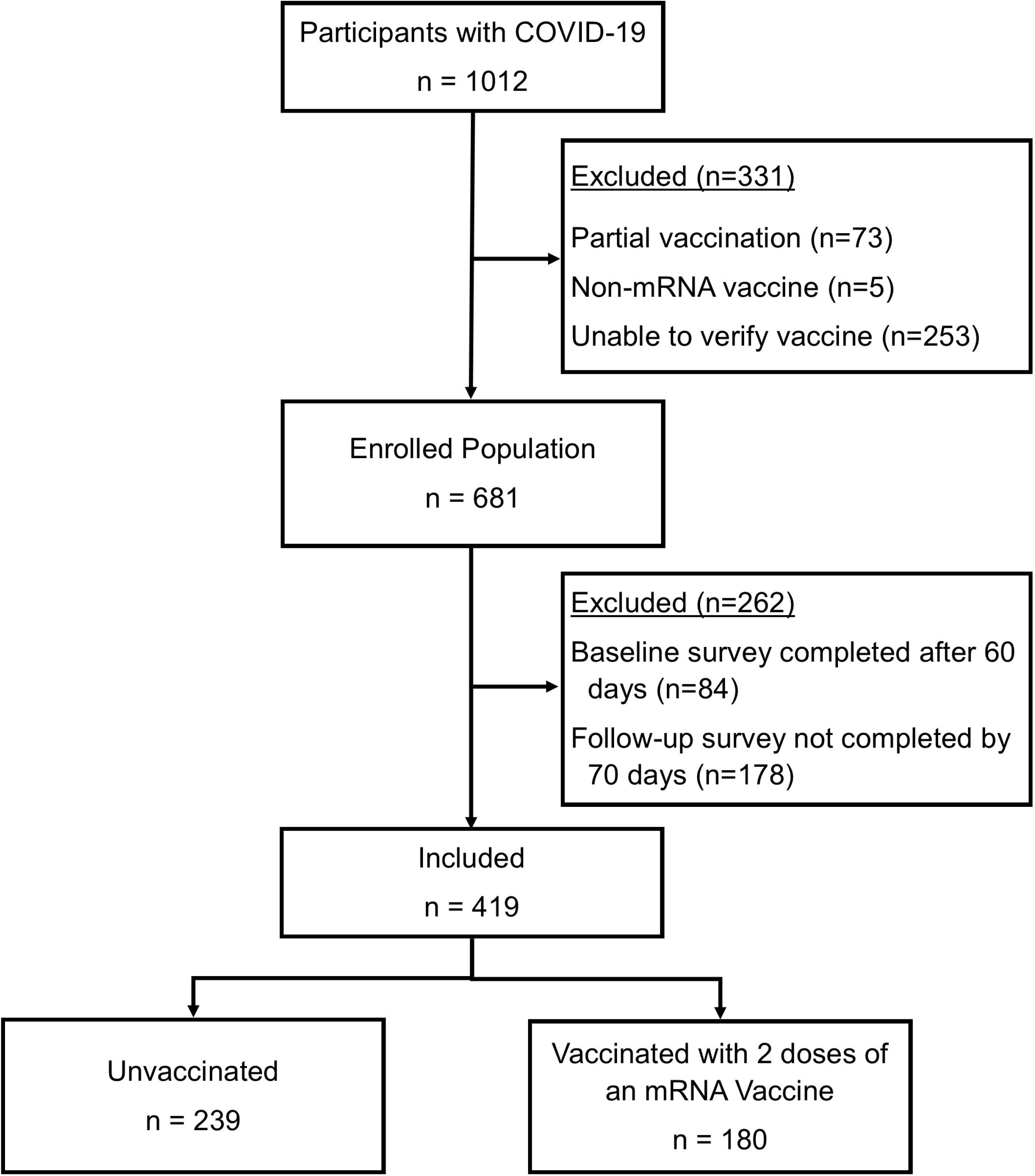
Enrollment of COVID-19 Vaccinated and Unvaccinated U.S. Healthcare Personnel.

**Table 1.** Demographic Characteristics and Comorbidities of Vaccinated and Unvaccinated U.S. Healthcare Personnel with COVID-19.

|  | All<br>Participants<br>(n=419)<br>n (%) | Vaccinated<br>(n=180)<br>n (%) | Not<br>Vaccinated<br>(n=239)<br>n (%) | p<sup>1</sup> |
| --- | --- | --- | --- | --- |
| **Age Group (Years)** |  |  |  | 0.019 |
| 18–29 | 90 (21.5) | 27 (15.0) | 63 (26.4) |  |
| 30–39 | 167 (39.9) | 73 (40.6) | 94 (39.3) |  |
| 40–49 | 85 (20.3) | 45 (25.0) | 40 (16.7) |  |
| 50–64 | 77 (18.4) | 35 (19.4) | 42 (17.6) |  |
| **Sex** |  |  |  | 0.342 |
| Male | 64 (15.3) | 32 (17.8) | 32 (13.4) |  |
| Female | 352 (84.0) | 146 (81.1) | 206 (86.2) |  |
| Non-binary | 1 (0.2) | 1 (0.6) | 0 |  |
| Missing data | 2 (0.5) | 1 (0.6) | 1 (0.4) |  |
| **Race and Ethnic Group** |  |  |  | <0.001 |
| White, non-Hispanic | 303 (72.3) | 145 (80.6) | 158 (66.1) |  |
| Black, non-Hispanic | 47 (11.2) | 9 (5.0) | 38 (15.9) |  |
| Hispanic or Latino | 41 (9.8) | 13 (7.2) | 28 (11.7) |  |
| Other, non-Hispanic | 28 (6.7) | 13 (7.2) | 15 (6.3) |  |
| **Education Level** |  |  |  | <0.001 |
| High school or less | 25 (6.0) | 6 (3.3) | 19 (8.0) |  |
| Undergraduate or technical degree | 293 (69.9) | 109 (60.6) | 184 (77.0) |  |
| Graduate or professional degree | 101 (24.1) | 65 (36.1) | 36 (15.1) |  |
| **Job Classification** |  |  |  | <0.001 |
| Non-clinical | 128 (30.5) | 46 (25.6) | 82 (34.3) |  |
| Physician | 20 (4.8) | 18 (10.0) | 2 (0.8) |  |
| Advanced Practice Provider | 12 (2.9) | 10 (5.6) | 2 (0.8) |  |
| Nurse/Nurse Assistant | 164 (39.1) | 57 (31.7) | 107 (44.8) |  |
| Housekeeping | 2 (0.5) | 0 | 2 (0.8) |  |
| Other Clinical | 48 (11.5) | 27 (15.0) | 21 (8.8) |  |
| Other | 45 (10.7) | 22 (12.2) | 23 (9.6) |  |
| **Health Insurance** |  |  |  | 0.004 |
| Private | 385 (91.9) | 175 (97.2) | 210 (87.9) |  |
| Government | 17 (4.1) | 3 (1.7) | 14 (5.9) |  |
| None | 3 (0.7) | 0 | 3 (1.3) |  |
| Unknown | 14 (3.3) | 2 (1.1) | 12 (5.0) |  |
| **Presence of 2 or more Comorbidities (see Supplemental Table 3 for Details)** | 216 (51.6) | 98 (54.4) | 118 (49.4) | 0.325 |

<sup>1</sup>Calculated using Fisher's exact test

Among participants included in the analysis, follow-up surveys were completed at a median of 6.0 weeks after first symptoms (IQR 6.0–6.3). Symptoms were common at 6 weeks, with 298 (71.1%) reporting at least one COVID-like symptom, 236 (56.3%) reporting at least one neurologic symptom, and 318 (75.9%) reporting any symptom (**Figure 2**). Among those with COVID-like symptoms at 6 weeks, 245 (95.7%) had symptoms that were also reported from the time of the initial diagnosis. Within 2 weeks of symptom onset, 323 (77.1%) participants had returned to work; by 6 weeks, only seven (1.7%) still had not returned.

**Figure 2.**
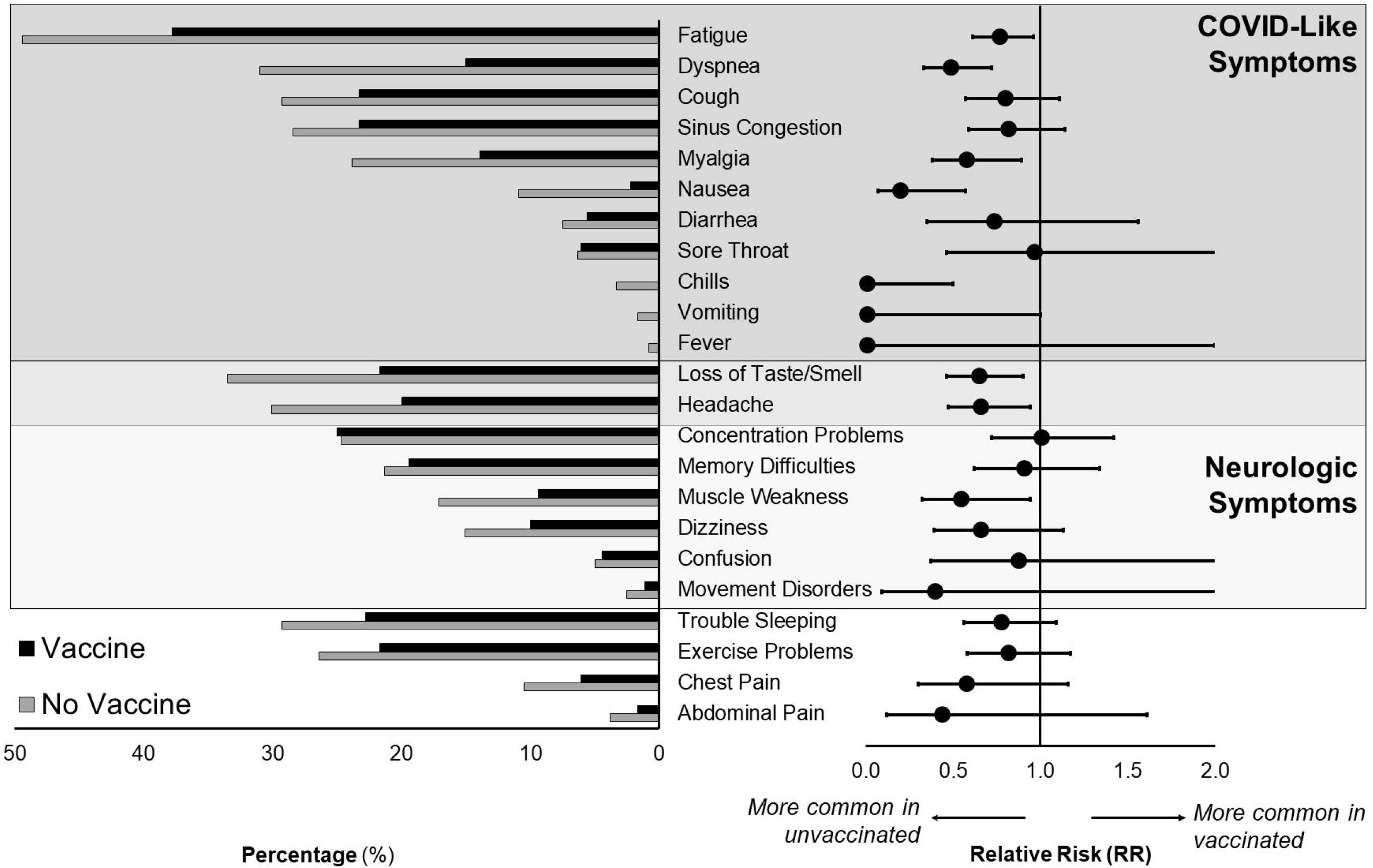
Prevalence of New or Persistent Symptoms 6 Weeks after COVID-19 Symptom Onset among U.S. Healthcare Personnel. Each bar in the left pane shows the percentage of participants reporting symptoms at the 6-week follow-up, stratified by vaccination status. For each symptom, the relative risk (RR, unadjusted) and 95% confidence interval are shown on the forest plot to the right. For RR <1.0, the symptom is less prevalent among the vaccinated. Note that several symptoms are part of both COVID-19 symptoms and neurologic symptoms. *COVID-like symptoms* included fever, cough, dyspnea, chills, fatigue, myalgia, headache, new loss of taste or smell, sore throat, nasal congestion, diarrhea, and nausea or vomiting. *Neurologic symptoms* included dizziness, headache, muscle weakness, movement disorders, confusion, memory difficulties, concentration problems, or loss of taste or smell. *Any symptoms* included trouble sleeping, exercise problems, chest pain, or abdominal pain, in addition to COVID-19 symptoms and neurologic symptoms, defined above. *RR, relative risk*.

Vaccinated participants had lower prevalence of COVID-like symptoms at the 6-week survey compared to those who were not vaccinated (60.6% vs. 79.1%). The relative risk for COVID-like symptoms was 0.77 (95% confidence interval [CI], 0.67–0.88) before adjustment and 0.70 (CI, 0.58–0.84) after adjustment for covariates. This risk ratio corresponds to an adjusted risk difference after 6 weeks of 24.1-percentage points (95% CI, 11.6–36.6 percentage points). Other classifications of symptoms were also less likely after vaccination—for neurologic symptoms the adjusted risk ratio (aRR) was 0.71 (95% CI 0.55–0.93) with a 17.9 percentage point reduction (95% CI 5.1–30.7); for any 6-week symptoms the aRR was 0.76 (95% CI 0.65– 0.90), with 20.1 percentage point reduction (95% CI 8.0–32.1; **Figure 3, Supplemental Table S6**).

**Figure 3.**
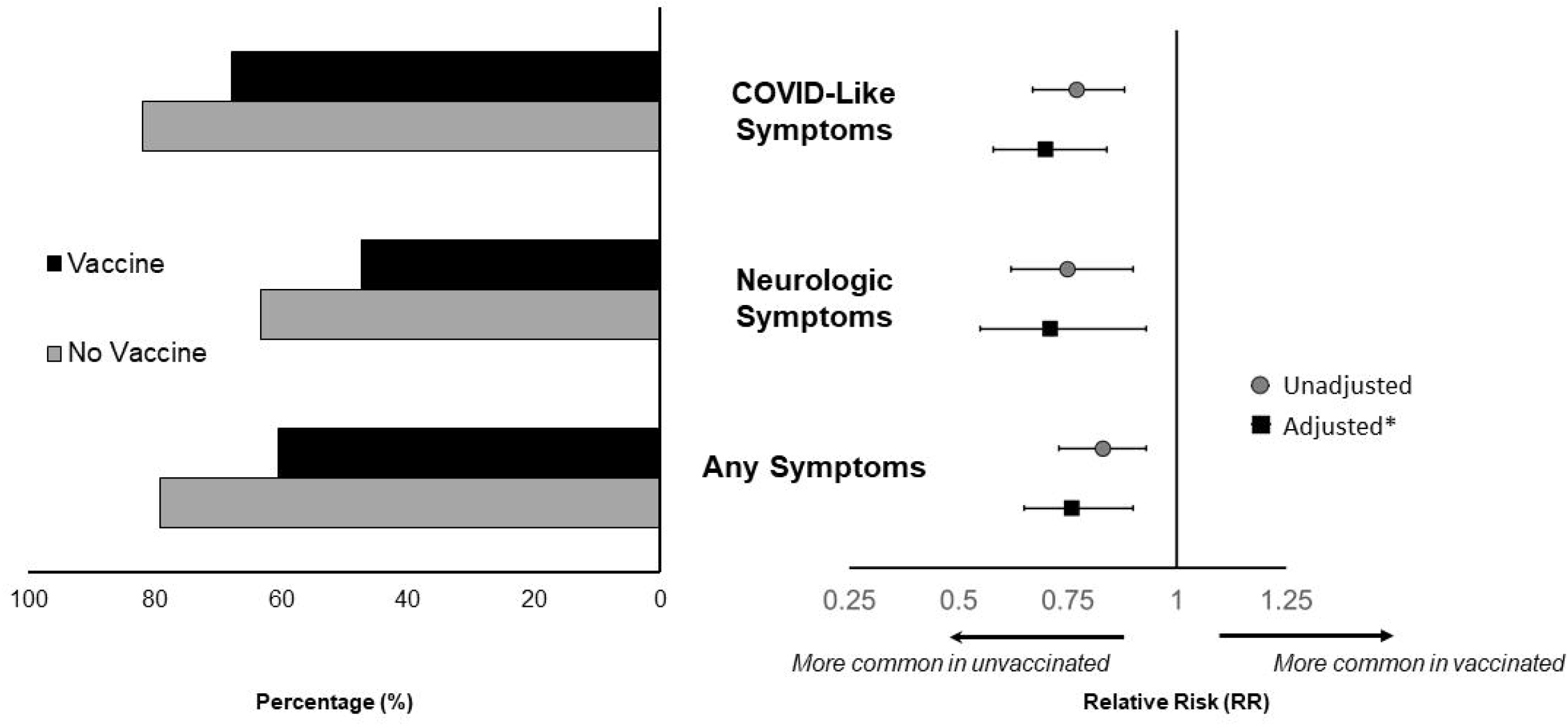
New or Persistent Symptoms at 6 Weeks after COVID-19 Symptom Onset in Vaccinated versus Unvaccinated U.S. Healthcare Personnel. Each bar in the left pane shows the percentage of participants reporting symptoms at the 6-week follow-up, stratified by vaccination status. This forest plot in the right pane shows the estimated risk of new or persistent symptoms present at the 6-week survey. The relative risk (RR) shows the ratio between the probability of having symptoms in the vaccinated vs. the unvaccinated (values <1.0 indicate that the prevalence of symptoms is lower in the vaccinated group than the unvaccinated). Grey circles show the unadjusted estimates, and black squares show the estimates adjusted for age, race, ethnicity, comorbidities, calendar month of diagnosis, and weeks since symptoms started. Error bars indicate 95% confidence intervals around the point estimate. *COVID-like symptoms* included fever, cough, dyspnea, chills, fatigue, myalgia, headache, new loss of taste or smell, sore throat, nasal congestion, diarrhea, and nausea or vomiting. *Neurologic symptoms* included dizziness, headache, muscle weakness, movement disorders, confusion, memory difficulties, concentration problems, or loss of taste or smell. *Any symptoms* included trouble sleeping, exercise problems, chest pain, or abdominal pain, in addition to COVID-19 symptoms and neurologic symptoms, defined above. * Adjusted for age, race, ethnicity, comorbidities, calendar month of diagnosis, and weeks since symptoms started.

Symptoms that were associated with being unvaccinated included dyspnea, myalgia, muscle weakness, fatigue, chills, loss of taste or smell, headache, and nausea; prevalence of other individual symptoms at 6 weeks did not differ significantly between vaccinated and unvaccinated participants (**Figure 2**). The sensitivity analysis restricted to persistent symptoms revealed similar findings (**Supplemental Figure S1**).

The median time from symptom onset to return to work was 13 days (IQR 11–16 days). Vaccinated participants returned to work a median of 2.0 days (95% CI 1.0–3.0) sooner than the unvaccinated and were less likely to return to work more than 10 days after illness onset (78.9% vs. 87.5%; RR 0.90; 95% CI 0.82–0.99). Adjusting for covariates, vaccinated participants returned to work sooner than unvaccinated participants (aHR, 1.37; 95% CI 1.04–1.79; **Supplemental Table S7)**. Vaccinated participants were also less likely to have COVID-like symptoms on return to work, although without statistical significance (49.4% vs 66.2%, RR 0.83; 95% CI 0.67–1.03). Participants who reported COVID-like symptoms on return to work were more likely than those without to report COVID-like symptoms at 6 weeks (84.7% vs 50.9%, RR 1.36; 95% CI 1.11–1.67). The time to return to work for each vaccination cohort is shown in **Figure 4**.

**Figure 4.**
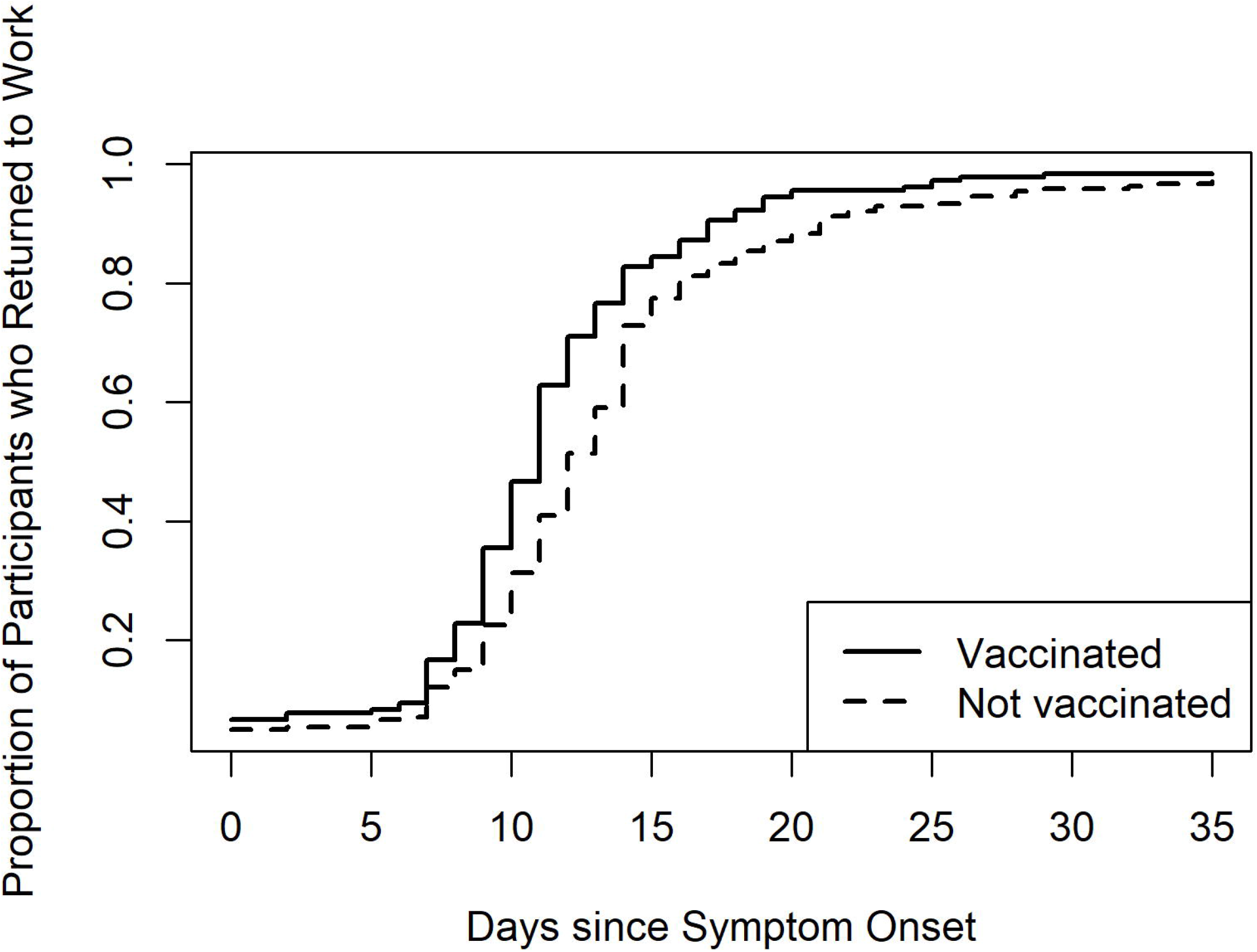
Kaplan-Meier Plot of Proportion of U.S. Healthcare Personnel Returning to Work after Onset of COVID-19 Symptoms, Stratified on COVID-19 Vaccination. The Kaplan-Meier plot shows the actual time to return to work, stratified on vaccination status (log-rank test, p <0.001). A Cox proportional hazards model was constructed, adjusting for age, race, ethnicity, comorbidities, and calendar month of diagnosis. The adjusted hazards ratio (aHR) for the adjusted model is 1.37 (95% CI 1.04 to 1.79). Note that aHR>1.0 indicates that participants *resume work* more quickly.

## Discussion

In this study of HCP diagnosed with COVID-19 between December 2020 and August 2021, we observed that 71% of participants with a confirmed diagnosis of COVID-19 had at least one COVID-like symptom at 6 weeks after symptom onset; nearly 76% had COVID-like or other symptoms, such as neurologic symptoms. This high proportion of symptoms suggests that HCP might be experiencing significant persistent disease burden. We observed that COVID-19 infection after full vaccination (breakthrough infection) was associated with a 24-percentage point absolute risk reduction of symptoms at 6 weeks compared with COVID-19 in unvaccinated HCP. Previous reports from the same study have estimated that COVID-19 vaccination is ≥88% effective at preventing COVID-19.^3^ Reduced likelihood of experiencing prolonged symptoms after COVID-19 is an additional benefit of vaccination and indicates that vaccine effectiveness against developing COVID-19 with prolonged symptoms is higher than vaccine effectiveness against COVID-19 alone. Together with evidence for vaccine effectiveness, our findings indicate that vaccination prevents prolonged symptoms both by preventing infection in the first place, and by hastening recovery if infection occurs.

Furthermore, vaccinated participants also returned to work 2 days sooner than non-vaccinated participants; it appears that vaccinated HCP were able to return to work sooner after infection due to fewer ongoing symptoms. Any effect, magnified over 22 million HCP in the U.S., is in addition to the benefit of vaccines in preventing infections, and can impact health system capacity and the ability to respond to public health emergencies.^33^ Similar effects might be expected among vaccinated employees in other critical industries.

While the effectiveness of COVID-19 vaccination at preventing infection and hospitalization is well described, the effect on duration of symptoms following infection is less clear. The UK-based COVID Symptom Study using self-reported data from a mobile app-based data collection instrument with unvalidated vaccination status and COVID-19 diagnosis found that any of 32 symptoms lasting ≥28 days was less prevalent among those who were vaccinated with two doses (aOR 0.51, 95% CI 0.32–0.82).^16^ This effect size is similar to findings in our study. The validation of testing and vaccination status in our defined HCP cohort are strengths of the current investigation.

We found that several specific symptoms 6 weeks after illness onset were most strongly associated with having no prior vaccination, including nausea, dyspnea, muscle weakness, myalgia, loss of taste or smell, and headache. Other symptoms had point estimates in the direction of vaccine effectiveness, even if the magnitude of effect did not reach statistical significance. These findings are important because neurologic and other symptoms are frequently reported several months after COVID-19.^13^ The differential association with vaccination of these symptoms might provide insight into their pathophysiology. Lower frequency of these symptoms following vaccination could be associated with decreased severity of initial illness, as vaccination is known to decrease severity of disease, and prior studies have found that prolonged symptoms might be more common among those with severe COVID-19 illness.^18,28^ Since an effect of vaccines in preventing prolonged symptoms is likely to be mediated by the immune response, further research is needed to understand the mechanisms of prolonged symptoms that might be amenable to other prevention strategies.

Our study has several limitations. First, our follow-up was limited to 6 weeks after symptom onset. While our survey response was robust and we validated test results and vaccinations, the prevalence of symptoms is likely to decay over time and we did not assess longer term effects. We were also not able to determine whether symptoms were caused by SARS-CoV-2 infection, although other evidence indicates that symptoms are usually specific to COVID-19 if reported 6 weeks after illness onset.^9^ Second, we relied on self-reported symptoms in participants who knew their vaccination status. Many reported symptoms are subjective, leaving open the possibility that vaccinated participants had more confidence that their symptoms would resolve quickly. It is also possible that symptom severity affected response rates, although our findings were similar after excluding symptoms that were rated as mild by participants. Third, our study period precluded analysis of booster doses, which are now recommended for adults in the United States.^34^ It is possible that booster doses might provide additional protection against symptoms after initial COVID-19. Our analysis was also limited to infections before introduction of the Omicron variant. Finally, there could be unmeasured differences between those vaccinated and unvaccinated that predispose some participants to having persistent symptoms or returning to work more quickly. We attempted to account for some of these factors in our regression models, and sensitivity analyses yielded similar findings to our primary analysis, but residual or unmeasured confounding could still have impacted our results.

In conclusion, a primary series of COVID-19 vaccination was associated with decreased risk of having new or persistent symptoms at 6 weeks and returning to work sooner in a cohort of HCP. Future work is warranted to assess the effect of vaccination on longer term symptoms, daily function, quality of life, and the effect in other populations.

## Data Availability

All data produced in the present study are available upon reasonable request to the authors.

## Acknowledgements

The authors would like to acknowledge the following participating Project PREVENT medical centers: Baystate Medical Center, Springfield, Massachusetts; Brigham and Women’s Hospital, Boston, Massachusetts; Jackson Memorial Hospital, Miami, Florida; University Medical Center New Orleans, LSU Health Sciences Center New Orleans, Louisiana; Olive View-University of California Los Angeles Medical Center, Los Angeles, California; Jefferson Health, Philadelphia, Pennsylvania; University Health-Truman Medical Center/University of Missouri-Kansas City, Kansas City, Missouri; University of Chicago, Chicago, Illinois; University of Iowa, Iowa City, Iowa; University of Massachusetts Chan Medical School, Worcester, Massachusetts; University of Mississippi Medical Center, Jackson, Mississippi; University of Alabama at Birmingham, Birmingham, Alabama; Community Medical Centers, Fresno, California; University of Washington, Seattle, Washington; and Valleywise Health Medical Center, Phoenix, Arizona.

The authors would also like to acknowledge the following individuals: Allison Schuette; Brianna M. DiFronzo; Karen Hopcia; Theresa M. Orechia; Alexander B. Hill; Gabrielle Donohoe; Lily R. Johnsky; Jordyn M. Fofi; Steven E. Miyawaki; Jenson J. Kaithamattam; Michelle Chung; Nikita A. Umale; Mohammad Adrian Hasdianda; Guruprasad Jambaulikar; Tala Teymour; Maria Davila; Suzette Fernandez; Joshua Tiao; Alexandria Henderson; Reynaldo Padilla; Cynthia Delgado; Madeleine Manahan; Melanie Potts; Jessica Kuo; Alyssa Fowlds; Zoe Speight; Laurie Kemble; Danielle Beckham; Lori Wilkerson; Geneatra Green; Rachel Marrs; Katherine Schneider; Cathy Fairfield; Fred Ullrich; Virginia Mangolds; Morgan Nelson; Abigail Lopes; Scott Pelletier; Gloria Essien; Rebekah Peacock; Alan Jones; Bhagyashri Navalkele; Savannah Vann; Andrea William; Brooke Park; Eugene Melvin; Joel Rodgers; Nivedita Patkar; Delissa Tidwell-Hand; Whitney Covington; Michael C. Kurz; Peter Poerzgen; Layla A. Anderson; Kyle A. Steinbock; Jennifer Smith; Amy Dakos; Denise Tritt; Stacey Wisniewski; Gaynell Bernadas-Hunt; Christine D. Crider; Susana Hacopian; Vincent E. Yu; Lidia Choxom; and Nathan R. Kramer.

