## Supplemental Tables and Figures for "Presence of Symptoms 6 Weeks After COVID-19 Among Vaccinated and Unvaccinated U.S. Healthcare Personnel"

| **TABLES** | **PAGE** |
| --- | --- |
| **Table S1.** Characteristics of Participating Medical Centers | 2 |
| **Table S2.** Qualifying Symptoms and Definitions | 3 |
| **Table S3.** Definition of Underlying Conditions for Analysis of Persistent Symptoms by Vaccination Status | 4 |
| **Table S4.** Demographic and Employment Factors of U.S. Healthcare Personnel Who Completed the 6-Week Survey Versus Those Who Did Not | 5 |
| **Table S5.** Comorbidities of U.S. Healthcare Personnel | 6 |
| **Table S6.** Relative Risk of Symptoms at 6 Weeks after COVID-19 Symptom Onset among U.S. Healthcare Personnel ≥14 days After Two Doses of an mRNA Vaccination Compared with No Vaccination | 7 |
| **Table S7.** Cox Proportional Hazards Model for Time to Return to Work among U.S. Healthcare Personnel | 8 |
| **Table S8.** STROBE Statement—Checklist | 9 |
| **FIGURES** |  |
| **Figure S1.** Prevalence of Persistent Symptoms 6 Weeks after COVID-19 Symptom Onset among U.S. Healthcare Personnel | 18 |

**Supplemental Table S1. Characteristics of Participating Medical Centers.**

|  | **Medical Centers**  **(n=15)** |
| --- | --- |
| **Total employees, median (IQR)** | 12,761 (10,544-16,750) |
| **Region, n (%)** |  |
| **Northeast** | 4 (27) |
| **Southeast** | 4 (27) |
| **Midwest** | 3 (20) |
| **West** | 4 (27) |
| **Academic medical center, n (%)** | 15 (100) |
| **Services, n (%)** |  |
| **Acute care hospital** | 15 (100) |
| **Long-term care facility** | 2 (13) |
| **Urgent Care** | 11 (73) |
| **Vaccination coverage in Sept 2021 (%), median (IQR)** | 84 (73-91) |

**Supplemental Table S2.** **Qualifying Symptoms and Definitions.** This table details the definitions for qualifying possible symptoms of COVID-19 that qualified for inclusion in the study cohort and for analysis of which symptoms were present at 6 weeks.

| **Symptoms** | **Enrollment symptoms^1^ (within 14 days of positive SARS-CoV-2 test)** | **COVID-like symptoms (at 6 weeks)** | **Neurologic symptoms (at 6 weeks)** | **Any 6-week symptoms (at 6 weeks)** |
| --- | --- | --- | --- | --- |
| Abdominal pain | X |  |  | X |
| Bruised toes or feet | X |  |  |  |
| Chest pain | X |  |  | X |
| Chills | X | X |  | X |
| Concentration problems |  |  | X | X |
| Confusion |  |  | X | X |
| Cough | X | X |  | X |
| Diarrhea | X | X |  | X |
| Difficulty with exercise |  |  |  | X |
| Dizziness |  |  | X | X |
| Fatigue | X | X |  | X |
| Fever | X | X |  | X |
| Headache | X | X | X | X |
| Loss of appetite | X |  |  |  |
| Loss of taste or smell | X | X | X | X |
| Memory difficulties |  |  | X | X |
| Movement problems |  |  | X | X |
| Muscle weakness |  | X | X | X |
| Nausea, or vomiting | X | X |  | X |
| Persistent joint pains or muscle aches | X | X |  | X |
| Rigors | X |  |  |  |
| Runny nose | X |  |  |  |
| Severe respiratory illness | X |  |  |  |
| Shortness of breath | X | X |  | X |
| Sinus congestion | X | X |  | X |
| Sore throat | X | X |  | X |
| Trouble sleeping |  |  |  | X |

^1^Included in the enrollment survey. Some terms differed in the enrollment survey: ‘chest pain or tightness’ was asked instead of ‘chest pain’; fever was specified as temperature ≥37.8°C or subjective fever; ‘altered taste or smell’ was asked instead of ‘loss of taste or smell’; ‘muscle aches’ was asked instead of ‘persistent joint pains or muscle aches’, and ‘sinus or nasal congestion’ was asked instead of ‘sinus congestion’. These terms were considered equivalent for purposes of analysis of which 6-week symptoms were ‘persistent’.

**Supplemental Table S3: Definition of Underlying Conditions for Analysis of Persistent Symptoms by Vaccination Status.**

This table details the recorded comorbidities (from which participants were dichotomized as having none versus 2 or greater).

Participants were considered to have two or more comorbidities if at least two conditions were present from the following list of conditions verified by self-report or the medical record:

- Active cancer;

- Alcohol use disorder;

- Allergic rhinitis;

- Anxiety/obsessive-compulsive/trauma or stressor related disorder;

- Asthma;

- Autoimmune rheumatologic disease;

- Chronic kidney disease;

- Chronic liver disease;

- Cognitive/neurodevelopmental disorder;

- COPD/emphysema;

- Coronary artery disease;

- Deep vein thrombosis or pulmonary embolism;

- Depression or other mood disorder;

- Diabetes mellitus;

- Dialysis;

- Hematopoietic stem cell transplant;

- Hypertension;

- Movement or motor disorders;

- Other chronic lung disease;

- Other heart condition;

- Other immunosuppressing condition;

- Other mental health condition;

- Sleep disorder;

- Solid organ transplant; or

- Stroke.

**Supplemental Table S4. Demographic and Employment Factors of U.S. Healthcare Personnel Who Completed the 6-Week Survey Versus Those Who Did Not**

|  | Completed 6-Week Survey^1^  (n=603)  n (%) | Did Not Complete 6-Week Survey^2^  (n=141)  n (%) | *p^3^* |
| --- | --- | --- | --- |
| **Age** |  |  | 0.2684 |
| 18-29 y | 90 (21.5) | 69 (26.4) |  |
| 30-39 y | 167 (39.9) | 86 (33.0) |  |
| 40-49 y | 85 (20.3) | 56 (21.5) |  |
| 50-64 y | 77 (18.4) | 50 (19.2) |  |
| **Sex** |  |  | 0.5491 |
| Male | 64 (15.4) | 43 (16.7) |  |
| Female | 352 (84.4) | 214 (83.0) |  |
| Non-binary | 1 (0.2) | 1 (0.3) |  |
| **Race and Ethnic Group** |  |  | 0.0072 |
| White, non-Hispanic | 303 (72.3) | 157 (60.2) |  |
| Black, non-Hispanic | 47 (11.2) | 35 (13.4) |  |
| Hispanic or Latino | 41 (9.8) | 40 (15.3) |  |
| Other, non-Hispanic | 28 (6.7) | 29 (11.1) |  |
| **Education Level** |  |  | 0.6669 |
| High school or less | 25 (6.0) | 15 (5.9) |  |
| Undergraduate or technical degree | 293 (69.9) | 187 (73.1) |  |
| Graduate or professional degree | 101 (24.1) | 54 (21.1) |  |
| **Job Classification** |  |  | 0.3525 |
| Non-clinical | 128 (30.6) | 71 (27.2) |  |
| Physician | 20 (4.8) | 13 (5.0) |  |
| Advanced Practice Provider | 12 (2.9) | 7 (2.7) |  |
| Nurse/Nurse Assistant | 128 (30.6) | 71 (27.2) |  |
| Housekeeping | 2 (0.5) | 0 |  |
| Other Clinical | 48 (11.5) | 20 (7.7) |  |
| Other | 45 (10.7) | 26 (10.0) |  |
| **Health Insurance** |  |  | 0.0123 |
| Private | 385 (91.9) | 220 (84.3) |  |
| Government | 17 (4.1) | 18 (6.9) |  |
| None | 3 (0.7) | 2 (0.8) |  |
| Unknown | 14 (3.3) | 21 (8.1) |  |
| **Presence of 2 or more Comorbidities** (see Supplemental Table 3 for Details) | 207 (49.4) | 107 (41.0) | 0.0331 |
| **Received Any COVID-19 Vaccine**^2^ | 192 (45.9) | 122 (46.7) | 0.0604 |

^1^ Completed follow-up survey with 10 weeks of index test. Those completing after 10 weeks are excluded from this analysis.

^2^ For those who did not complete follow-up survey, vaccine status is determined by self-report.

^3^ Calculated using Fisher’s exact test

**Supplemental Table S5. Comorbidities of** **U.S. Healthcare Personnel.** This table compares the comorbidities between vaccinated and unvaccinated participants.

|  | All Participants  (n=419)  n (%) | Vaccinated  (n=180)  n (%) | Not Vaccinated  (n=239)  n (%) | *p*^1^ |
| --- | --- | --- | --- | --- |
| **Comorbidities** |  |  |  |  |
| Asthma | 49 (11.7) | 19 (10.6) | 30 (12.6) | 0.544 |
| Allergic Rhinitis | 32 (7.6) | 19 (10.6) | 13 (5.4) | 0.063 |
| COPD/emphysema | 0 | 0 | 0 | 0.999 |
| Other Chronic Lung Disease | 3 (0.7) | 3 (1.7) | 0 | 0.079 |
| Hypertension | 63 (15.0) | 29 (16.1) | 34 (14.2) | 0.679 |
| Coronary Artery Disease | 1 (0.2) | 1 (0.6) | 0 | 0.430 |
| Other Heart Condition | 9 (2.1) | 7 (3.9) | 2 (0.8) | 0.042 |
| Stroke | 0 | 0 | 0 | 0.999 |
| Diabetes | 25 (6.0) | 12 (6.7) | 13 (5.4) | 0.679 |
| Chronic Kidney Disease | 0 | 0 | 0 | 0.999 |
| Dialysis | 1 (0.2) | 0 | 1 (0.4) | 0.999 |
| Solid Organ Transplant | 1 (0.2) | 0 | 1 (0.4) | 0.999 |
| Hematopoietic Stem Cell Transplant | 0 | 0 | 0 | 0.999 |
| Autoimmune Rheumatologic Disease | 16 (3.8) | 9 (5.0) | 7 (2.9) | 0.310 |
| Other Immunosuppressing Condition | 3 (0.7) | 1 (0.6) | 2 (0.8) | 0.999 |
| Active Cancer | 2 (0.5) | 0 | 2 (0.8) | 0.509 |
| Deep Vein Thrombosis or Pulmonary Embolism | 3 (0.7) | 2 (1.1) | 1 (0.4) | 0.579 |
| Chronic Liver Disease | 1 (0.2) | 1 (0.6) | 0 | 0.430 |
| Depression or Other Mood Disorder | 53 (12.6) | 31 (17.2) | 22 (9.2) | 0.017 |
| Anxiety/Obsessive-Compulsive/Trauma or Stressor Related Disorder | 63 (15.0) | 32 (17.8) | 31 (13.0) | 0.214 |
| Other Mental Health Condition | 3 (0.7) | 2 (1.1) | 1 (0.4) | 0.579 |
| Movement or Motor Disorders | 1 (0.2) | 1 (0.6) | 0 | 0.430 |
| Alcohol use disorder | 0 | 0 | 0 | 0.999 |
| Sleep Disorder | 21 (5.0) | 12 (6.7) | 9 (3.8) | 0.184 |
| Cognitive/Neurodevelopmental Disorder | 2 (0.5) | 2 (1.1) | 0 | 0.184 |

^1^Calculated using Fisher’s exact test

**Supplemental Table S6. Relative Risk of Symptoms at 6 Weeks after COVID-19 Symptom Onset among U.S. Healthcare Personnel ≥14 days After Two Doses of an mRNA Vaccination Compared with No Vaccination.**

|  | **Presence of Symptoms** | | | **Moderate Symptoms or Worse^†^** | |
| --- | --- | --- | --- | --- | --- |
| **Symptoms** | **Unadjusted RR (95% CI)** | **Adjusted RR (95% CI)** | **Adjusted Absolute Risk Difference% (95% CI)** | **Unadjusted RR (95% CI)** | **Adjusted RR (95% CI)** |
| COVID-like Symptoms | 0.77  (0.67–0.88) | 0.70  (0.58–0.84) | -24.1  ([-36.6]–[-11.6]) | 0.69  (0.52–0.93) | 0.67  (0.44–1.02) |
| Neurologic Symptoms | 0.75  (0.62–.90) | 0.71  (0.55–0.93) | -17.9  ([-30.7]–[-5.1]) | 0.58  (0.39–0.86) | 0.43  (0.26–0.73) |
| Any 6-week Symptoms | 0.83  (0.73–0.93) | 0.76  (0.65–0.90) | -20.1  ([-32.1]–[-8.0]) | 0.72  (0.55–0.95) | 0.70  (0.48–1.03) |

*COVID-19 symptoms* included fever, cough, dyspnea, chills, fatigue, myalgia, headache, loss of taste or smell, sore throat, nasal congestion or rhinorrhea, diarrhea, and nausea or vomiting. *Neurologic symptoms* included dizziness, headache, muscle weakness, movement disorders, confusion, memory difficulties, concentration problems, or loss of taste or smell. *Any symptoms* included fatigue, sinus congestion, trouble sleeping, exercise problems, chest pain, nausea, or abdominal pain, in addition to COVID-19 symptoms and neurologic symptoms, defined above.

^†^ Severity of illness was classified for each symptom by self-report into mild, moderate, or severe. “Moderate symptoms or worse” includes all symptoms rated as moderate or severe.

*RR, relative risk; 95% CI, 95% confidence interval*

**Supplemental Table S7. Cox Proportional Hazards Model for Time to Return to Work among U.S. Healthcare Personnel.** The outcome of this model is return to work, so hazard ratios (HR)>1.0 indicate a factor is associated with returning to work sooner.

|  | **Unadjusted HR (95% CI)** | **Adjusted HR (95% CI)** |
| --- | --- | --- |
| Vaccination Status |  |  |
| No Vaccination | Ref | Ref |
| Full Vaccination | 1.401 (1.151–1.704) | 1.367 (1.043–1.793) |
| Calendar Month |  |  |
| December | – | Ref |
| January | – | 1.477 (1.002–2.176) |
| February | – | 1.165 (0.712–1.908) |
| March | – | 0.973 (0.582–1.625) |
| April | – | 1.287 (0.765–2.167) |
| May | – | 1.231 (0.706–2.145) |
| June | – | 1.324 (0.746–2.351) |
| July | – | 1.331 (0.856–2.068) |
| August | – | 1.363 (0.582–2.181) |
| Age group (years) |  |  |
| 18–29 | – | Ref |
| 30–39 | – | 1.113 (0.854–1.450) |
| 40–49 | – | 1.024 (0.749–1.401) |
| 50–64 | – | 0.912 (0.662–1.256) |
| Race/Ethnicity |  |  |
| White, non-Hispanic | – | Ref |
| Black, non-Hispanic | – | 0.877 (0.637–1.206) |
| Hispanic or Latino | – | 0.742 (0.597–1.187) |
| Other, non-Hispanic | – | 0.867 (0.578–1.303) |
| More than 2 comorbid risk factors | – | 0.802 (0.654–0.982) |

**Supplemental Table S8: STROBE Statement—Checklist.**

|  | | | Item No. | | Recommendation | | | Page  No. | | | Relevant text from manuscript | |
| --- | --- | --- | --- | --- | --- | --- | --- | --- | --- | --- | --- | --- |
| **Title and abstract** | | | 1 | | (*a*) Indicate the study’s design with a commonly used term in the title or the abstract | | | 1 | | | Presence of Symptoms 6 Weeks After COVID-19 Among Vaccinated and Unvaccinated U.S. Healthcare Personnel | |
|  |  |  |  |  | (*b*) Provide in the abstract an informative and balanced summary of what was done and what was found | | | 3 | | | A history of COVID-19 mRNA vaccination among U.S. HCP with COVID-19 illness was associated with decreased risk of COVID-like symptoms at 6 weeks and earlier to return to work. | |
| Introduction | | | | | | | | | | | |  |
| Background/rationale | | | 2 | | Explain the scientific background and rationale for the investigation being reported | | 6 | | | Vaccination may prevent prolonged COVID-19 symptoms through several mechanisms: preventing COVID-19 infection , limiting the severity of acute illness in vaccine breakthrough cases through vaccine-mediated immunity, and affecting the ongoing immune response even after acute infection. | | |
| Objectives | | | 3 | | State specific objectives, including any prespecified hypotheses | | 7 | | | In this study our primary objective was to measure the association between prior mRNA COVID-19 vaccination and symptoms 6 weeks after acute COVID-19 infection among healthcare personnel (HCP). | | |
| Methods | | | | | | | | | | | |  |
| Study design | | | 4 | Present key elements of study design early in the paper | | 7 | | For this nested cohort study… | | | | |
| Setting | | | 5 | Describe the setting, locations, and relevant dates, including periods of recruitment, exposure, follow-up, and data collection | | 7 | | Characteristics of the 15 participating sites are summarized in Supplemental Table S1, and details of study protocols and forms for the parent study are available online.19 | | | | |
| Participants | | | 6 | (*a*) *Cohort study*—Give the eligibility criteria, and the sources and methods of selection of participants. Describe methods of follow-up  *Case-control study*—Give the eligibility criteria, and the sources and methods of case ascertainment and control selection. Give the rationale for the choice of cases and controls  *Cross-sectional study*—Give the eligibility criteria, and the sources and methods of selection of participants | | 7 | | we included HCP if they had a positive SARS-CoV-2 nucleic acid amplification test or antigen test and had symptoms consistent with COVID-19 (as listed in Supplemental Table S2) within 14 days of the positive test… Each participant completed an enrollment survey 14 to 60 days after his/her positive test, and was offered a follow-up survey 6 weeks after symptom onset. | | | | |
|  |  |  |  | (*b*) *Cohort study*—For matched studies, give matching criteria and number of exposed and unexposed  *Case-control study*—For matched studies, give matching criteria and the number of controls per case | |  | |  | | | | |
| Variables | | | 7 | Clearly define all outcomes, exposures, predictors, potential confounders, and effect modifiers. Give diagnostic criteria, if applicable | | 8 | | The 6-week survey included questions on a variety of symptoms which we categorized into three overlapping groups… | | | | |
| Data sources/ measurement | | | 8* | For each variable of interest, give sources of data and details of methods of assessment (measurement). Describe comparability of assessment methods if there is more than one group | | 9 | | As part of the follow-up survey we asked participants to report the dates when they left and resumed work. We calculated time to return to work as the number of days from onset of symptoms until the first day at work after illness. | | | | |
| Bias | | | 9 | Describe any efforts to address potential sources of bias | | 10 | | We included in our multivariable model categorical variables of age, race, ethnicity, and a dichotomous variable for two or more comorbidities at baseline. | | | | |
| Study size | | | 10 | Explain how the study size was arrived at | | 7 | | As part of the PReventing Emerging Infections through Vaccine EffectiveNess Testing Project (Project PREVENT), we enrolled HCP who were working on-site at participating academic medical centers who had been tested for SARS-CoV-2 | | | | |
| Quantitative variables | | 11 | | Explain how quantitative variables were handled in the analyses. If applicable, describe which groupings were chosen and why | | 10 | | We used a Cox proportional hazards model to calculate an adjusted hazard ratio (aHR) to compare time to return to work between fully vaccinated and unvaccinated participants, counting zero days if there were no days off work after symptom onset. | | | | |
| Statistical methods | | 12 | | (*a*) Describe all statistical methods, including those used to control for confounding | | 10-11 | | We used multivariable Poisson regression with a sandwich variance estimator to predict the relative risk of having symptoms at the 6-week follow-up for complete vaccination compared with no vaccination | | | | |
|  |  |  |  | (*b*) Describe any methods used to examine subgroups and interactions | | 9 | | We defined our primary outcome as the presence of COVID-like symptoms at the time of the 6-week survey. We conducted additional analyses for neurologic symptoms and for any 6-week symptoms, and we assessed whether symptoms at 6 weeks were also present within 14 days of the date of the positive test | | | | |
|  |  |  |  | (*c*) Explain how missing data were addressed | | 11 | | We included 419 HCP who were diagnosed with COVID-19 between December 28, 2020 and August 26, 2021, including 180 (43.0%) who were fully vaccinated with an mRNA vaccine and 239 (57.0%) who were not vaccinated; 41 were excluded because of unverified vaccination status and 143 were partially vaccinated (Figure 1). | | | | |
|  |  |  |  | (*d*) *Cohort study*—If applicable, explain how loss to follow-up was addressed  *Case-control study*—If applicable, explain how matching of cases and controls was addressed  *Cross-sectional study*—If applicable, describe analytical methods taking account of sampling strategy | | 11 | | Those who did not complete the follow-up survey had fewer comorbidities and were less likely to be White non-Hispanic (Supplemental Table S4). | | | | |
|  |  |  |  | (*e*) Describe any sensitivity analyses | | 9 | | For assessment of all symptom groups we performed a sensitivity analysis restricted to symptoms rated by participants as “moderate” or “severe”. | | | | |
| Results | | | | | | | | | | | | |
| Participants | | 13* | | | (a) Report numbers of individuals at each stage of study—eg numbers potentially eligible, examined for eligibility, confirmed eligible, included in the study, completing follow-up, and analysed | | | F1 | | | | See Figure 1 |
|  |  |  |  |  | (b) Give reasons for non-participation at each stage | | | F1 | | | | See Figure 1 |
|  |  |  |  |  | (c) Consider use of a flow diagram | | | F1 | | | | See Figure 1 |
| Descriptive data | | 14* | | | (a) Give characteristics of study participants (eg demographic, clinical, social) and information on exposures and potential confounders | | | 11 | | | | See Table 1, Table S5 |
|  |  |  |  |  | (b) Indicate number of participants with missing data for each variable of interest | | | T1 | | | | See Table 1 |
|  |  |  |  |  | (c) *Cohort study*—Summarise follow-up time (eg, average and total amount) | | | 11 | | | | Follow-up surveys were completed at a median of 6.0 weeks after first symptoms (IQR 6.0-6.3). |
| Outcome data | | 15* | | | *Cohort study*—Report numbers of outcome events or summary measures over time | | | 11 | | | | Symptoms were common at 6 weeks, with 298 (71.1%) reporting at least one COVID-like symptom, 236 (56.3%) reporting at least one neurologic symptom, and 318 (75.9%) reporting any symptom (Figure 2). |
|  |  |  |  |  | *Case-control study—*Report numbers in each exposure category, or summary measures of exposure | | |  | | | |  |
|  |  |  |  |  | *Cross-sectional study—*Report numbers of outcome events or summary measures | | |  | | | |  |
| Main results | | 16 | | | (*a*) Give unadjusted estimates and, if applicable, confounder-adjusted estimates and their precision (eg, 95% confidence interval). Make clear which confounders were adjusted for and why they were included | | | 11 | | | | The relative risk for COVID-like symptoms was 0.77 (95% confidence interval [CI], 0.67 to 0.88) before adjustment and 0.70 (CI, 0.58 to 0.84) after adjustment for covariates. |
|  |  |  |  |  | (*b*) Report category boundaries when continuous variables were categorized | | | T1 | | | | See Table 1 |
|  |  |  |  |  | (*c*) If relevant, consider translating estimates of relative risk into absolute risk for a meaningful time period | | | 11-12 | | | | This risk ratio corresponds with an adjusted risk difference after 6 weeks of 24.1-percentage points (95% CI, 11.6 to 36.6 percentage points). |
| Other analyses | 17 | | | | Report other analyses done—eg analyses of subgroups and interactions, and sensitivity analyses | | | | 12 | | | Neurologic or any 6-week symptoms after COVID-19 were also less prevalent among vaccinated compared with unvaccinated participants, and these findings were preserved restricting analysis to moderate or severe symptoms (Figure 3, Supplemental Table S6). |
| Discussion | | | | | | | | | | | | |
| Key results | 18 | | Summarise key results with reference to study objectives | | | | | 13 | | | | We observed that COVID-19 infection after full vaccination (breakthrough infection) was associated with a 24-percentage point absolute risk reduction of symptoms at 6 weeks compared with COVID-19 in unvaccinated HCP. |
| Limitations | 19 | | Discuss limitations of the study, taking into account sources of potential bias or imprecision. Discuss both direction and magnitude of any potential bias | | | | | 15 | | | | Our study has several limitations. First, our follow-up was limited to 6 weeks after symptom onset.. |
| Interpretation | 20 | | Give a cautious overall interpretation of results considering objectives, limitations, multiplicity of analyses, results from similar studies, and other relevant evidence | | | | | 16 | | | | In conclusion, COVID-19 vaccination was associated with decreased risk of having new or persistent symptoms at 6 weeks and returning to work sooner in a cohort of HCP. |
| Generalisability | 21 | | Discuss the generalisability (external validity) of the study results | | | | | 16 | | | | Future work is warranted to assess the effect of vaccination on longer term symptoms, daily function, quality of life, and the effect in those who do not work in healthcare. |
| Other information | | |  | | | | | | | | | |
| Funding | 22 | | Give the source of funding and the role of the funders for the present study and, if applicable, for the original study on which the present article is based | | | | | 1-2 | | | | This project was funded by the Centers for Disease Control and Prevention (CDC) (U01CK000480). The project is additionally supported by the Institute for Clinical and Translational Science at the University of Iowa through a grant from the National Center for Advancing Translational Sciences at the National Institutes of Health (UL1TR002537). |

*Give information separately for cases and controls in case-control studies and, if applicable, for exposed and unexposed groups in cohort and cross-sectional studies.

**Note:** An Explanation and Elaboration article discusses each checklist item and gives methodological background and published examples of transparent reporting. The STROBE checklist is best used in conjunction with this article (freely available on the Web sites of PLoS Medicine at http://www.plosmedicine.org/, Annals of Internal Medicine at http://www.annals.org/, and Epidemiology at http://www.epidem.com/). Information on the STROBE Initiative is available at www.strobe-statement.org.

**Supplemental Figure S1. Prevalence of Persistent Symptoms 6 Weeks after COVID-19 Symptom Onset among U.S. Healthcare Personnel.** Compared with Figure 2, this figure removes symptoms that developed between 2 and 6 weeks. Each bar in the left pane reports the percentage of participants reporting symptoms at the 6-week follow-up, stratified by vaccination status. For each symptom, the relative risk (RR) and 95% confidence interval are listed to the right of the bar. For RR <1.0, the symptom is more prevalent among the unvaccinated. Note that several symptoms are part of both COVID-19 symptoms and neurologic symptoms.

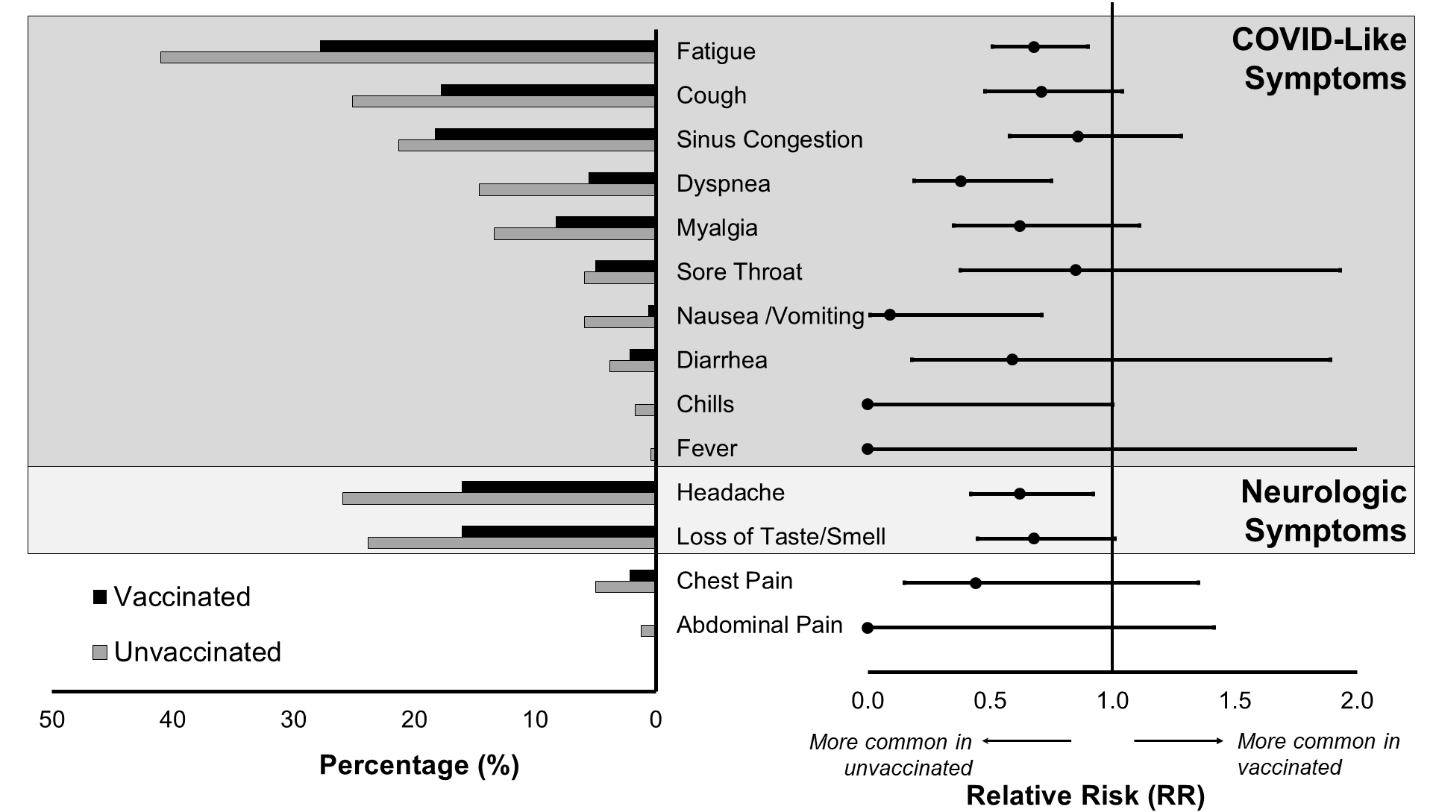

*RR, relative risk*
